# Less Wrong COVID-19 Projections With Interactive Assumptions

**DOI:** 10.1101/2020.06.06.20124495

**Authors:** Aditya Nagori, Raghav Awasthi, Vineet Joshi, Suryatej Reddy Vyalla, Akhil Jarodia, Chandan Gupta, Amogh Gulati, Harsh Bandhey, Keerat Kaur Guliani, Mehrab Singh Gill, Ponnurangam Kumaraguru, Tavpritesh Sethi

## Abstract

COVID-19 pandemic is an enigma with uncertainty caused by multiple biological and health systems factors. Although many models have been developed all around the world, transparent models that allow interacting with the assumptions will become more important as we test various strategies for lockdown, testing and social interventions and enable effective policy decisions. In this paper, we developed a suite of models to guide the development of policies under different scenarios when the lockdown opens. These had been deployed to create an interactive dashboard called *COVision* which includes the Agent-based Models (ABM) and classical compartmental models (CCM). Our tool allows simulation of scenarios by changing the strength of lockdown, basic reproduction number(R0), asymptomatic spread, testing rate, contact rate, recovery rate, incubation period, leakage in lockdown etc. We optimized ABM and CCMs and evaluated them on multiple error metrics. Out of these models in our suite, ABM was able to capture the data better than CCMs. Our evaluation suggests that ABM models were able to capture the dynamic nature of the epidemic for a longer duration of time while CCMs performed inefficiently. We computed R0 using CCMs which were found to be decreasing with lockdown duration, indicating the effectiveness of policies in different states of India. Models have been deployed on a dashboard hosted at http://covision.tavlab.iiitd.edu.in which allows users to simulate outcomes under different parameters and will allow the policymakers to make informed decisions and efficient monitoring of the covid19 pandemic in India.

## Introduction

The recent novel coronavirus disease, COVID-19, has shaken the world by storm. A communicable disease with no existing medical treatment, the World Health Organisation (WHO) has already declared it a pandemic. Many countries have adopted the social distancing norms to limit the spread of this virus and are trying their best to contain the virus by nationwide lockdowns, testing, marking disease hotspots and contact tracing. Similarly, India had also implemented a 21-day lockdown up to 15th April 2020 to assess the situation, which was later extended to 3rd May 2020. Since prolonged lockdowns, including the closing of organizations, schools and educational institutions and other important places can cause depression in the economy and may cause mayhem among people in shortage of necessary living supplies, including food, shelter and hygienic environments, it is both necessary and imperative to model the flow of the infection so that the policymakers can assess the situation and consequently take constructive steps to save lives as well as the economy.

Since the start of the epidemic, many new studies and papers have come to the surface, aiming to model the spread of the disease in many countries, including India. While many have tried to give projections using machine and deep learning techniques, these tend to overfit the data [1] and are unable to accommodate the overtime changes in the parameters such as contact rate due to government interventions, such as lockdowns, increased testing, etc. However, most of the papers have used classical population-level epidemiological models, mainly Susceptible - Infected - Recovered (SIR) model, and some other models which are just some minor modifications of the same [1-2]. Even though these models are reliable for modern modelling of an epidemic or pandemic, they cannot contain the changes in the parameters, as is the case with the machine learning techniques. Even though the researchers have tried to take the effects of social distancing and lockdown while setting some parameters like contact rate or reproduction number, the numbers predicted using such dynamical models have not been reliable, as in the case of [3], which projects a sharp decrease after 25th March, which was not the case.

In this paper, we found that the classical epidemiological models cannot be used to model the disease effectively and reliably and promote the applications of agent-based models (ABMs). Many studies had used agent-based models instead of equation-based models in the past to study epidemics. Venkataraman et al. [4] argued against using compartmental models as they fail to capture individual-level dynamics as well as social heterogeneity in real-world epidemics. Instead, they used an agent-based modelling mechanism for forecasting the 2014-2015 Ebola epidemic which generated close to real-world scenarios for longer-term forecasts. Tuomisto et al. [5] used ABMs to identify potentially destructive policies that may have been undertaken by the government due to the shortcomings as explained accompanying various model strategies and their underlying assumptions.

## METHODS

### 1. Dataset

Datasets were obtained from the ministry of health and family welfare website[6]. The statewise data was downloaded on a daily basis and collated in a time-series format. Data were in the cumulative form; we extracted the infected counts by subtracting the cured/recovered and migrated counts from the total to get the infected counts. Exposure counts were obtained by taking the infected time series at a lag of the incubation period. Population and population density data were taken from the official Indian government websites[7,8]. The population data is used to represent susceptible populations in the models. The population density was taken from census 2020 of India. We had done our projection modelling in every state of India but here we are presenting the results of the top 6 states which were most affected till 20th May (Supplementary Figure 1).

### 2. Agent-Based Model

The agent-based model was used to predict the statewise daily and total new cases by simulating the spread on a population of agents. The agent-based model is built on top of an SEIR model with the entire population divided among four primary states of susceptible, exposed, infected & recovered. In order to incorporate the effect of interventions like lockdown and testing rate, we also introduced two additional states, ‘Under Lockdown’ and ‘Tested & Quarantined’. On initialization, a specified number of agents were randomly spread over an area with few agents exposed to the virus and the rest in a susceptible state. After an incubation period, the earlier exposed agents move to the infected state and start infecting more based on the provided contact rate value. The infected agents were further symptomatic & asymptomatic in nature, with the symptomatic agent having higher chances of being tested and isolated. We also introduced the concept of leakage in lockdown which is used to cover cases like religious gathering, movement of migrant workers and many others. Figure 1 shows the model’s state chart that guides an agent’s behaviour based on the state they were currently in.

**Figure 1.**
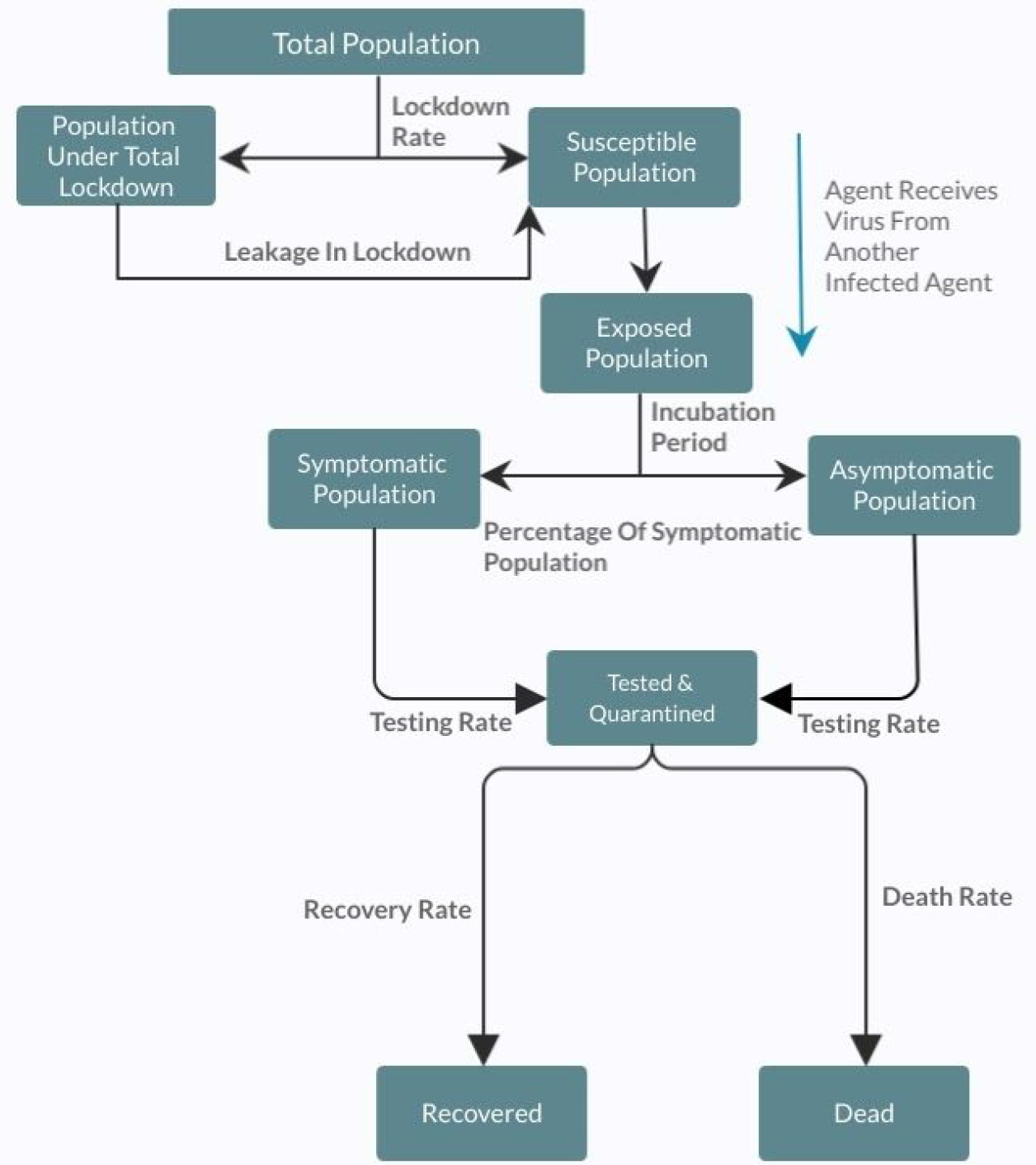
Flowchart showing the possible states of an agent and effect of interventions on them.

The ABM was developed in Anylogic, a multi-agent simulation software [9], because of its ability to allow highly customizable and scalable models.

Different combinations of parameters were evaluated using R^2^, Root Mean Squared Error (RMSE) and Mean Absolute Percentage Error (MAPE) of Predicted new cases vs actual new cases. The optimal model was selected from a grid search of combinations of parameters for future projections (Supplementary Table 1).

Data-Driven Contact Rates Scaling: To assemble the demographic variation of states in our models, we assumed the contact rate as a nonlinear function of population density and estimated state-wise contact rate using a Non-Linear Transmission Scaling Function proposed by Hao et al [10] (Supplementary Text3). In this nonlinear function, we assumed characteristic density constant (ρ_o_) as the density of Infection (Total Infected Count /Total Area under consideration) and r_max_ as the maximum distance of physical activity.

### 3. Classical Compartmental models & their optimisation

We used the standard compartmental ecological models called susceptible-infection-recovery (SIR) and susceptible-exposure-infection-recovery (SEIR). The model parameters involved the Contact rate (beta), Recovery rate (gamma) and Delta were optimised using ODE Solver and squared error minimisation. We numerically solved the ordinary differential equations in the SIR & SEIR (Supplementary text4) using the Runge-Kutta method as a first-order ordinary differential equation [11]. Assumptions for model parameters for recovery rate were taken from COVID literature; we found that the average time taken to recover by an individual with non-chronic illness is 2-3 weeks and for a chronic illness individual 3-6 weeks as mentioned in the WHO report [12]. We, therefore, used the range to optimise the recovery rate. The incubation period was taken in the range of 1-14. The exposure time-series were taken at a lag of the incubation period.

### 4. Dashboard

Scenarios: A user-friendly, interactive dashboard is developed in order to allow policymakers, researchers and the general public to get a better sense of the situation on a daily basis. We have currently incorporated optimised ABM and SIR, SEIR models along with various parameters. These models are optimised and updated with new data and user inputs in fractions of seconds.

Interactivity: The dashboard allows users to set R 0, Recovery rate and incubation period in literature-backed range for classical compartmental models. For ABM, the dashboard provides a variety of simulated scenarios including the best fitted and the Unlock models. These scenarios provide a sense on the spread of the disease and help in studying the effects of interventions. “Under the hood” option is added to read more about the methods used to produce these scenarios. The interface is shown in supplementary Figure 4.

### 5. Hospital beds reporting

For ICU beds requirement forecasting, we assumed an average of 9% [13] infected people needed critical care which varies in the range of 6%-12% [14,15,16] and used projection outcome of infected count from different models.

## Results

### 1. Quantitative evaluation of Models

To identify the best suite of forecasting models we quantitatively evaluated the compartmental and Agent-based models in different scenarios using Rsq, RMSE and Mape. By looking at RMSE, we observed that in the longer duration ABM has better performance than compartmental models (Figure 2), but for the shorter duration compartmental (SIR and SEIR models) models had comparable results. R-squared of ABM models remained equal to or above 90% suggesting the ability to capture the future trend of the new cases with higher accuracy. Mean absolute percentage errors (Mape) also remained smaller than in SIR and SEIR models, implying a lower percentage of absolute error in the prediction of new cases.

**Figure 2:**
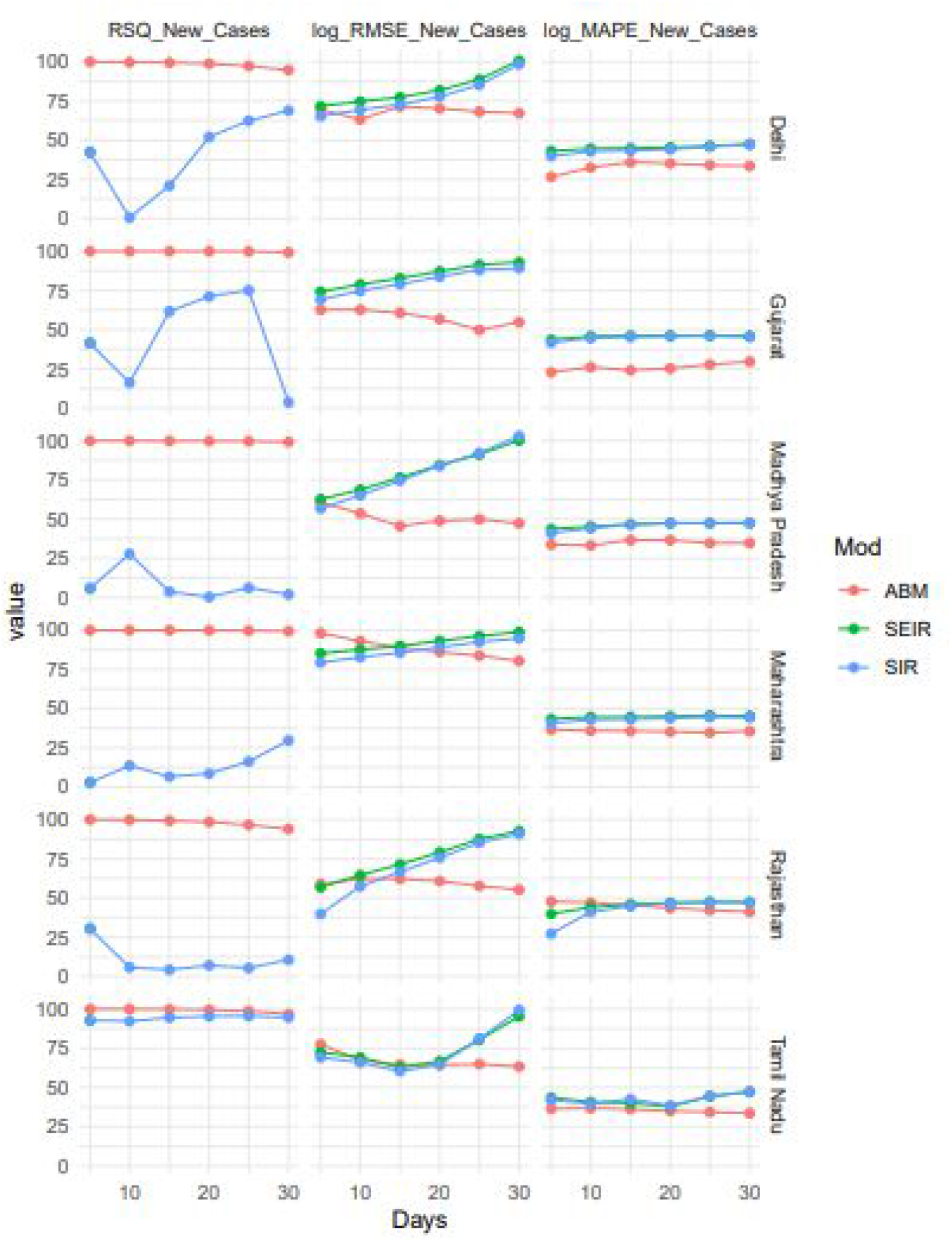
Comparison of State-wise projection from different models, state-wise R-squared, log Root Mean Square Error of daily new cases from different models, log Mean Absolute Percentage Error of daily new cases from different models are shown in the plots above. (Log base 1.1 is taken to match the scales for plotting) (Plot shows evaluation for Delhi, Gujrat, Madhya Pradesh, Maharashtra, Rajasthan, Tamil Nadu)

### 2. State-wise scenario-based (ABM) projections

Every state is different and holds geopolitical heterogeneity. The agent-based models (ABM) provide a variety of parameters that can all be tuned independently in order to capture the dynamic nature of the epidemic and state heterogeneity. Table 1 shows the list of all the parameters that are customizable in our model along with the values of all parameters that were kept the same for all the states. Parameters such as population, incubation rate, average illness duration, recovery rate, death rate, testing rate and per cent symptomatic[17] were taken from literature whereas the model was optimized around the fixed values of the area, initial infection and leakage in lockdown. Out of the remaining 2 parameters, lockdown rate was empirically changed to simulate the scenarios that provide the best fit and contact rate was calculated as described above.

**Table 1:** Scenarios/Parameters in Agent-Based Models.

| S.No. | Parameter | Description | Default Values |
| --- | --- | --- | --- |
| 1. | Population | Number of susceptible agents. | State-wise |
| 2. | Area | Dimension of the grid environment on which agents are spread. | Square root of population. |
| 3. | Incubation Period | Number of days an agent stays in the exposed state before moving to the infected one. | 7-14 days. |
| 4. | Percentage Symptomatic | Percentage of infected population showing symptoms. | 20% |
| 5. | Testing Rate | Percentage of agents tested. | State-wise |
| 6. | Contact Rate | Average number of agents an infected agent meets each day. | State-wise |
| 7. | Initial Infected | Number of infected agents before the model starts. | 2 |
| 8. | Lockdown Rate | Percentage of susceptible population that is under total lockdown. | State-wise |
| 10. | Leakage Rate | Percentage Of population moving from total lockdown to susceptible state. | 1% PM |
| 9. | Recovery Rate | Percentage of infected population that gets recovered. | State-wise |
| 10. | Mortality Rate | Percentage of infected population that dies. | State-wise |
| 11. | Average Illness Duration | Number of days after which an agent recovers or dies. | 21-42 days. |

**Table 2:**
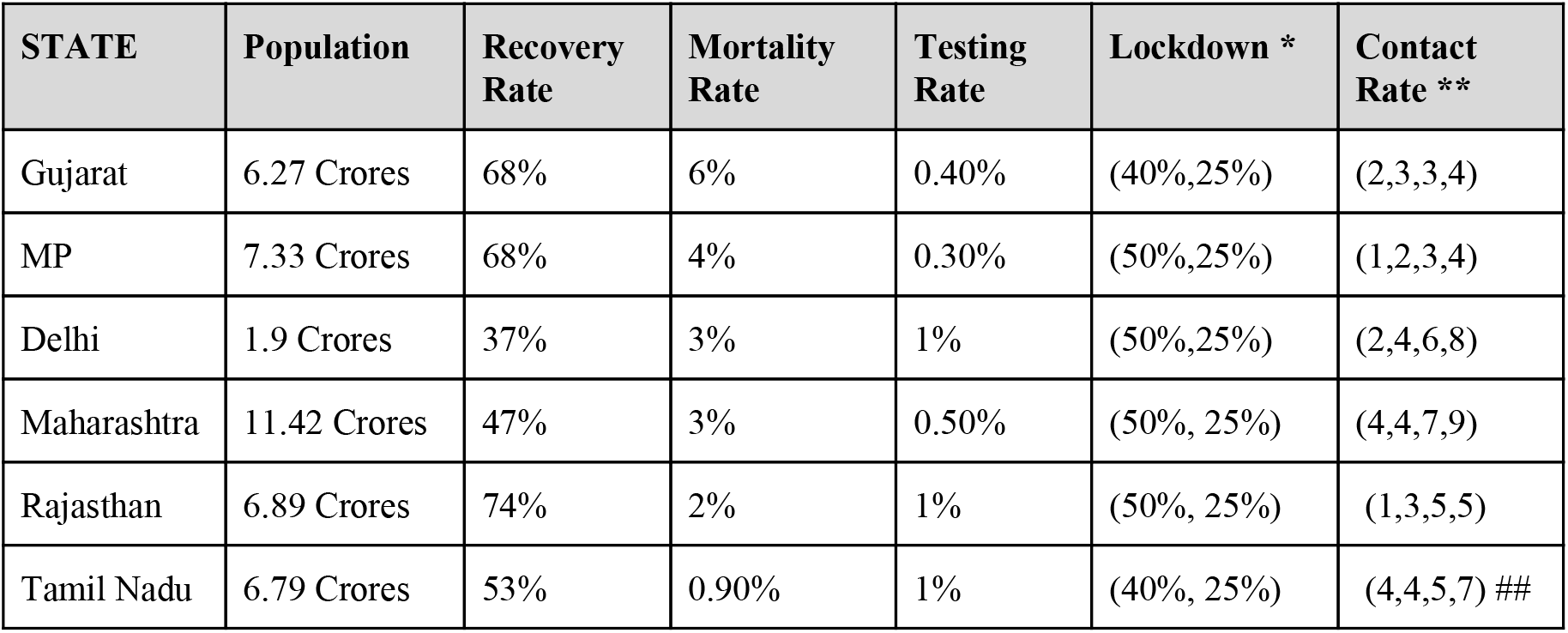
Best fitting scenarios for different states. * (Before 1st June, After 1st June) ** (Before 18th May, 18th May - 1st June, 1st June - 8th June, After 8th June) ## (Before 8th May,8th May - 1st June, 1st June - 8th June, After 8th June)

### 3. Unlock1: Lifting of the lockdown

The Indian government announced that from June 8th, 2020 [18], more services will be resumed in a planned manner. This is being done in lieu to subvert the economic crisis in the country. But this might increase the contact rate and hence the exposure of the larger percentage of the population to infection. We simulated these scenarios in ABM models for future predictions of new cases for the next 30 days. We observed a hockey stick point in the projections around the 5th to 8th June as shown in figure 3. These results were obtained by putting parameters the same as the best fit till 31th May, later than that, the lockdown was set to 25%, testing was taken from state-specific data, and contact rate was tuned according to population density and state-level government guidelines on restriction and relaxations. These simulations are available on our dashboard and can be tested for different parameters as well.

**Figure 3:**
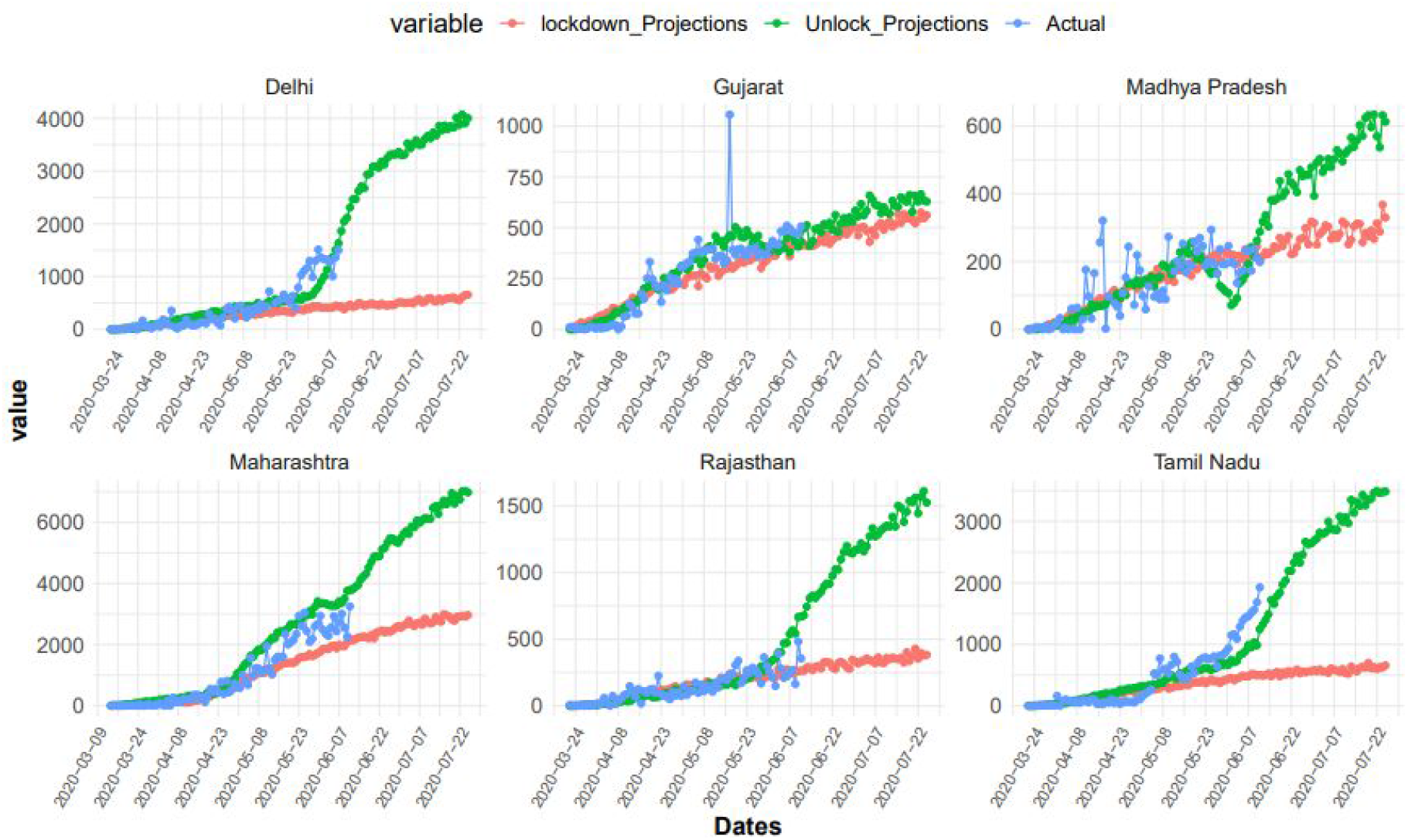
Unlock 1 and Lockdown differences of new cases using ABM shown for India, Delhi, Gujrat, Madhya Pradesh, Maharashtra, Rajasthan, Tamil Nadu.

### 4. Decreasing trend of R_0_ revealed the effectiveness of interventions

Since R_0_ is a good indicator to assess the policies and interventions, we trained our models on different stages of pandemic and calculated R_0_, we found that the trend of R_0_ was decreasing (See Figure 4) for every model, which suggests the effectiveness of the interventions. Initially at the smaller number of total cases, the fall in R_0_ was large as compared to when larger number of total cases were taken for R_0_ computation. Trend for Rajasthan and Gujrat was found to be ambiguous when compared between the SIR and SEIR models, however trend for India and others states followed a falling trend in R_0_.

**Figure 4:**
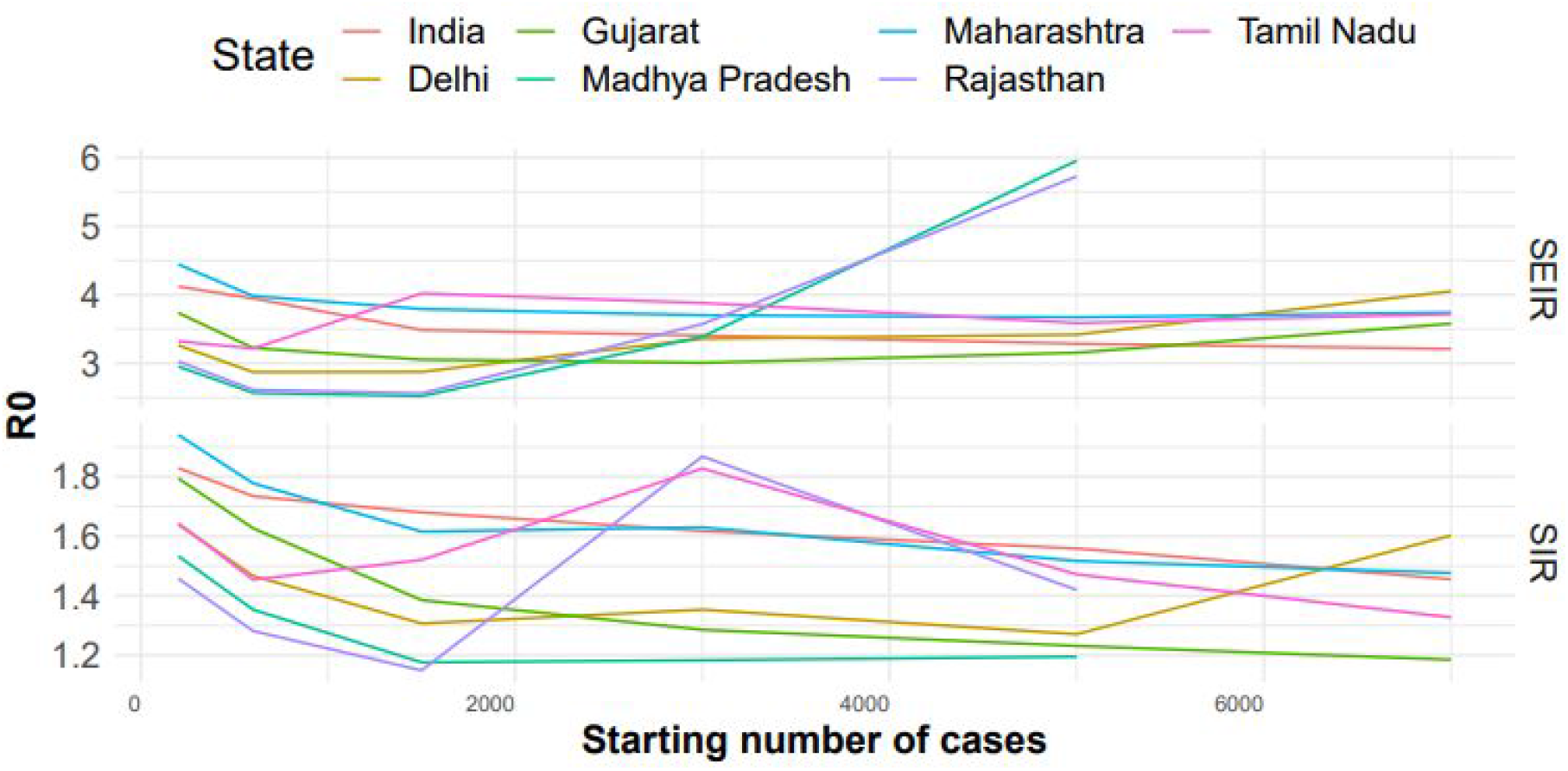
State-wise variation of R0 obtained from SIR and SEIR model for different periods of lockdown.

### 5. State-wise ICU Beds Forecasting

Assuming that 9% [6%,12%] of infected people require critical care, from the Agent-based model, we estimated the ICU beds requirement for every state from 11 june 2020 to 25 July 2020. We found that after 1st July Maharashtra will require more than 500 ICU Beds on a daily basis and after 29th June Delhi will require more than 300 ICU beds on a daily basis. For Gujarat and Madhya Pradesh, we found till 25th July, the maximum required is 56 and 55 per day respectively.

**Figure 5:**
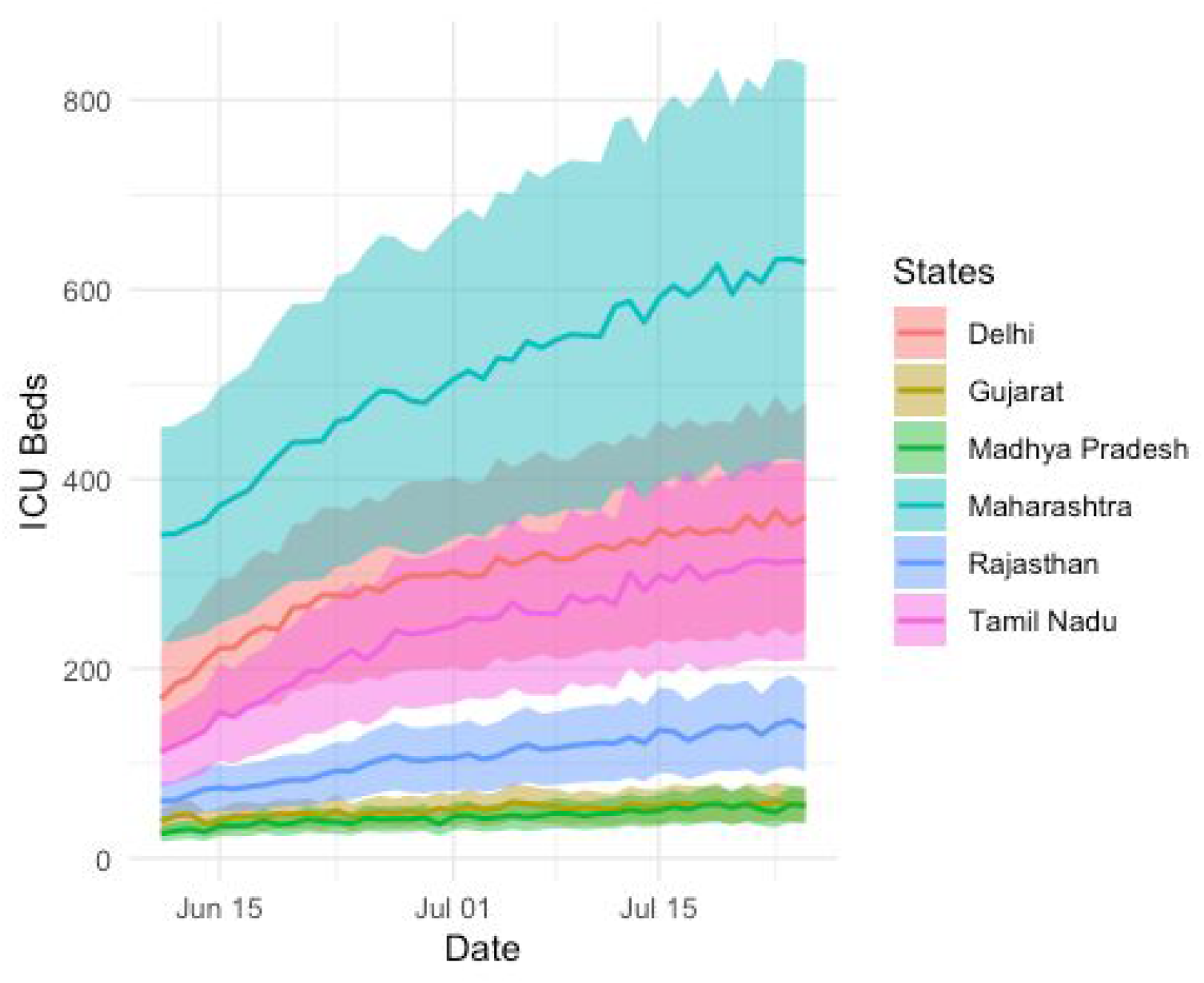
Daily ICU beds requirement forecasting from 11 June to 25 July using Agent-Based Model

## Discussion

Exponential spread of COVID-19 is becoming a threat to human survival and until no treatment is discovered social distancing is the only vaccine. India is the second most populated country in the world [19] and in high population density areas social distancing becomes difficult. One possible solution to tackle this is lockdown. Indian government implemented lockdown very timely and effectively and that controlled the spread of disease significantly. Since the spread of COVID is impacting multiple sectors like industry, education, agriculture, health and many more, effective interventions are required which need the projection of spread, in this study we have provided different models with different scenarios to project disease with an interactive dashboard. The ecological models were trained on the data downloaded from the Ministry of Health and Welfare website. Building these models involves parameter optimisation, a slight error in a given parameter can lead to error propagation in the final projections. We have therefore used minimum parameter optimisation. We did an extensive literature review to get a reasonable range of parameters. The WHO reports the recovery interval for healthy and chronic ill subjects as 2-3 weeks and 4-6 weeks respectively. [12]. The recovery rate in models were therefore optimised in this range. The incubation period has also been reported that it might take 1-14 [12] days to show the symptoms, thus, the incubation period of 1-14 days is a good guess for the optimisation. We had less clue about contact rates so we allowed our model to optimise in a wider range for the same. We looked at the literature and found out that our contact rates were in close proximity to what has been reported recently [2]. But these are classical models and making them robust by involving the dynamicity of human interventions and behaviours is little trickier, thus, we involved the Agent-based Models (ABM) which allowed us to incorporate the wider interventional measures. Lockdown in India had a gradual implementation as we saw migrant worker movement and religious gatherings increase the spread during the first lockdown starting from 24th March 2020. The ability of ABMs to customize interventions and incorporate such scenarios allows them to capture this behaviour in a better way and study how lockdown and testing might affect the upcoming trend. We have scaled contact rates used in ABM using population density and infection density. The function simulates the contact rate as population density increases, initially, the contact-rates rise as the contact combinations increases. However, beyond a certain point, the value of contact rate begins to stagnate as there are limits to individual movements and connections and hence any further increase in population density does not affect contact rate. Thus, the curve becomes stagnant at higher population density (supplementary text3).

Testing rate affects the asymptomatic population more as symptomatic agents are more likely to be tested in the first place. It should be noted that there are many models being currently studied and deployed related to COVID 19 in India and other countries. As noted by Holmdahl and Buckley [20], the selectivity for a particular model depends upon various factors, like the time period for the application, the underlying assumptions, and the parameters used and their estimations. It goes on to further argue to check for the specificity of the model, and identify the type of data used to fit the model. The most important parameter, the contact rate [20], needs to be correctly estimated and must take into account various lockdown and reopening scenarios if the model is to be used in the long run. Hence, it is extremely important to specify the assumptions that were made while constructing a model, which are able to accommodate for rapidly changing crises and select the model which complements our task.

## Data Availability

All data were used in the manuscript are publicly available from the Ministry of Health and Family Welfare, Government of India website.

## Acknowledgements

This work was partially supported by the Wellcome Trust/DBT India Alliance Fellowship IA/CPHE/14/1/501504 awarded to Dr. Tavpritesh Sethi and the Center for Artificial Intelligence at IIIT-Delhi. Authors also acknowledge Prof. Rakesh Lodha from All India Institute of Medical Sciences, New Delhi, for his valuable inputs. We also thank CSIR for supporting Mr Aditya Nagori and UGC to support Mr Raghav Awasthi with PhD fellowships. Special thanks to Ms Bhavika Rana for her design input for the dashboard frontend.

## Authors Contributions

Concept TP, PK, AN, RA Compartmental models: AN, RA, CG, TP ABM Models: VJ, AJ, TP Dashboard Frontend: SR, AG, BR, PK Dashboard Backend AN, VJ, AJ, HB Statistical Analysis: AN, RA, VJ Data Extraction & Curation: AN, HB Manuscript: AN, RA,VJ,CG,TP

## Supplementary Material

### 1. Top 6 states which were most affected till 20th May

**Supplementary Figure 1.**
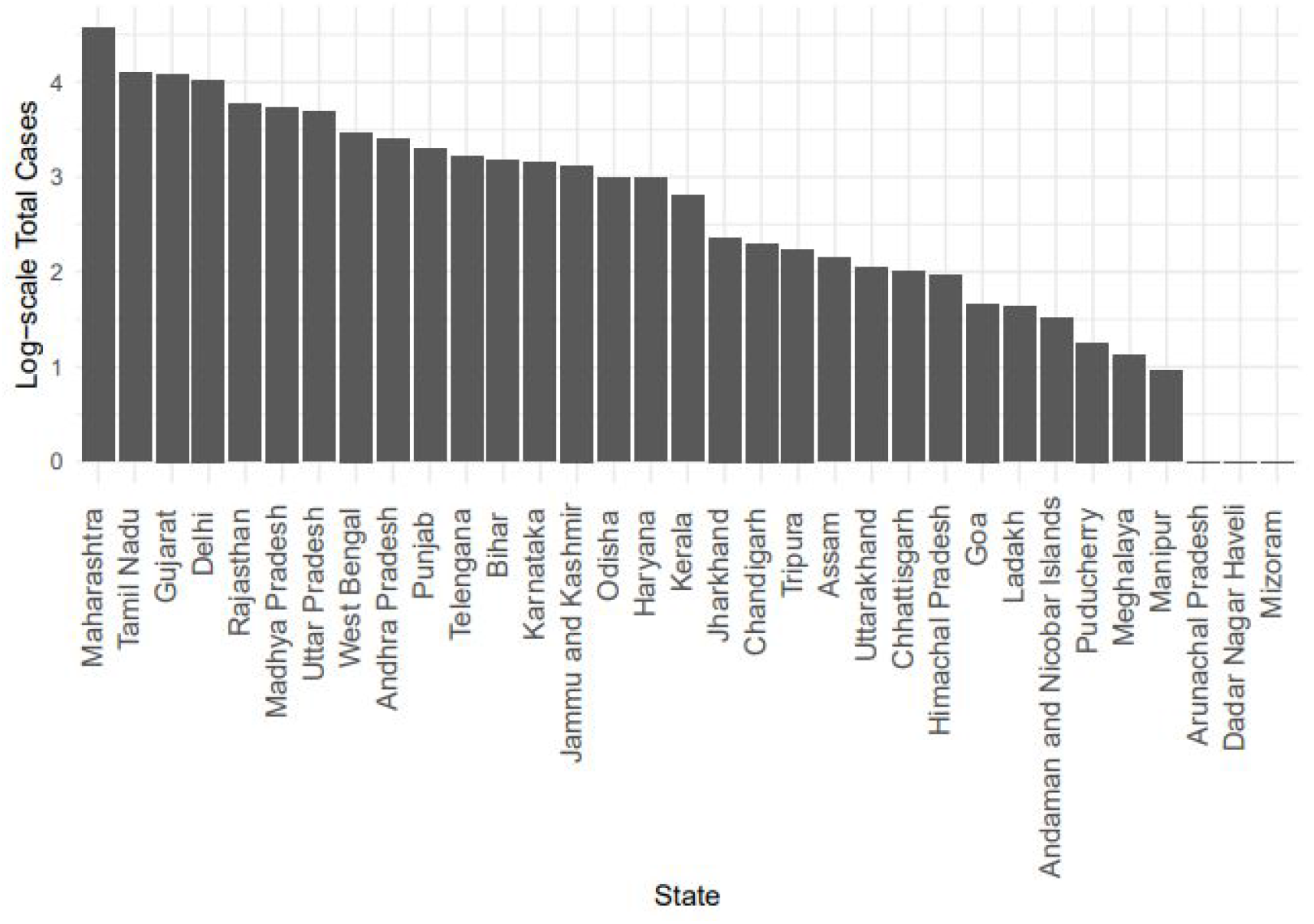
Barplot log_10_ of total number of COVID-19 cases until 20th May.

### 2. ABM Parameter selection

We run multiple simulations with different combinations of the parameters to ensure the best fit of the projected daily count to the actual cases (supplementary Figure 2). Different combinations which were tested are listed in supplementary table 1.

**Supplementary Figure 2:**
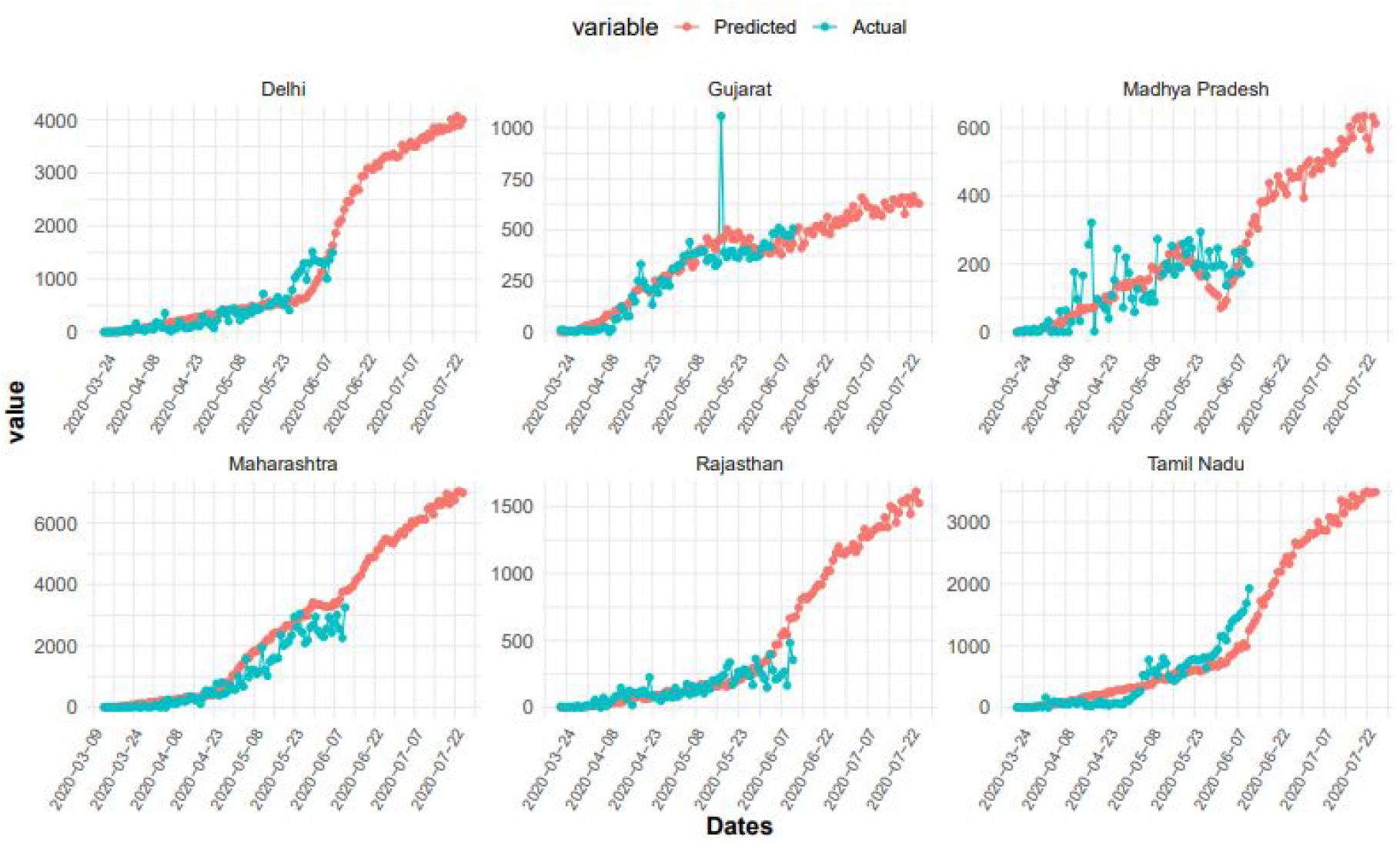
ABM based Projections of daily new cases in states i.e Delhi, Gujrat, Madhya Pradesh, Maharashtra, Rajasthan, Tamil Nadu

**Supplementary Table 1.**
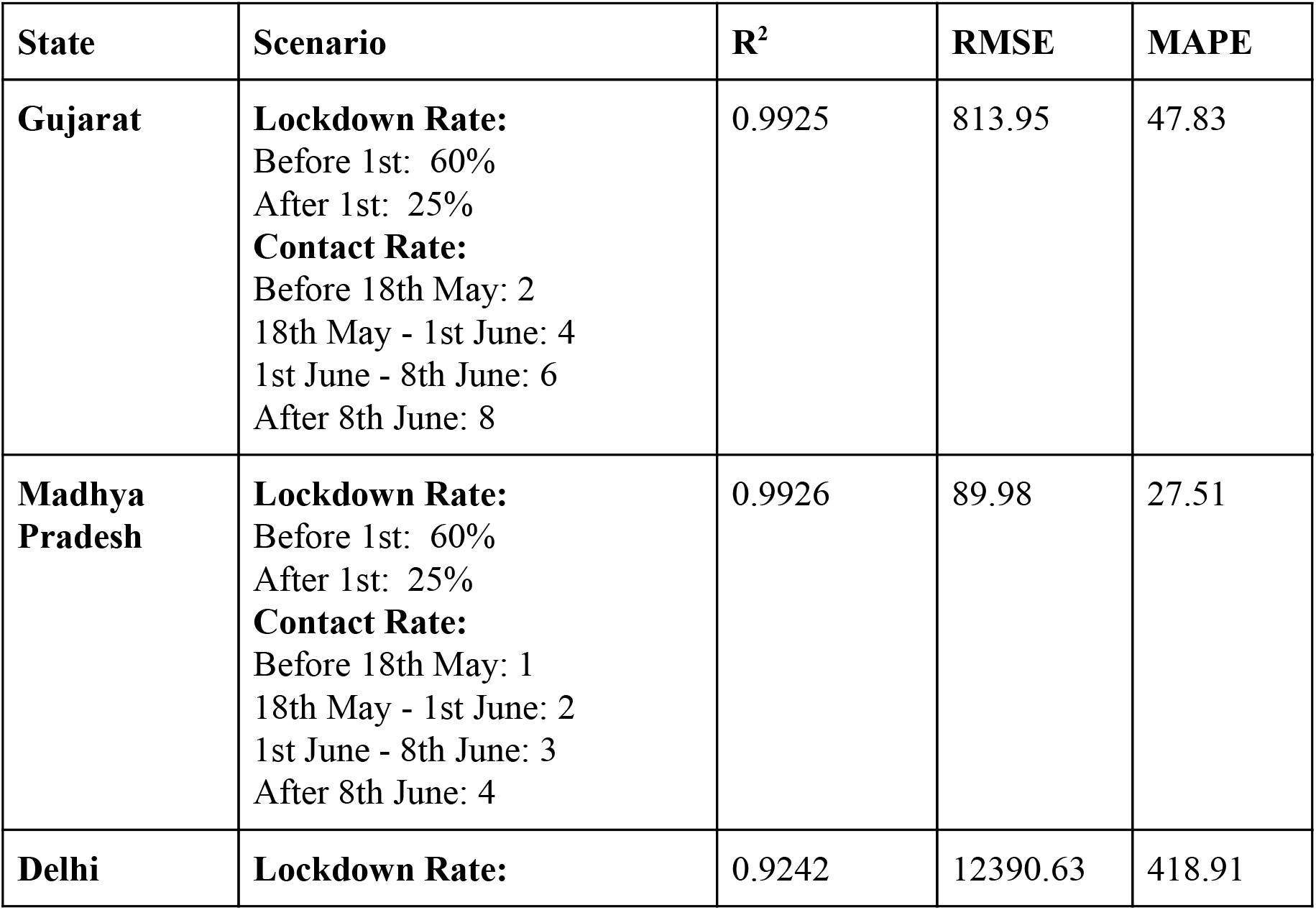
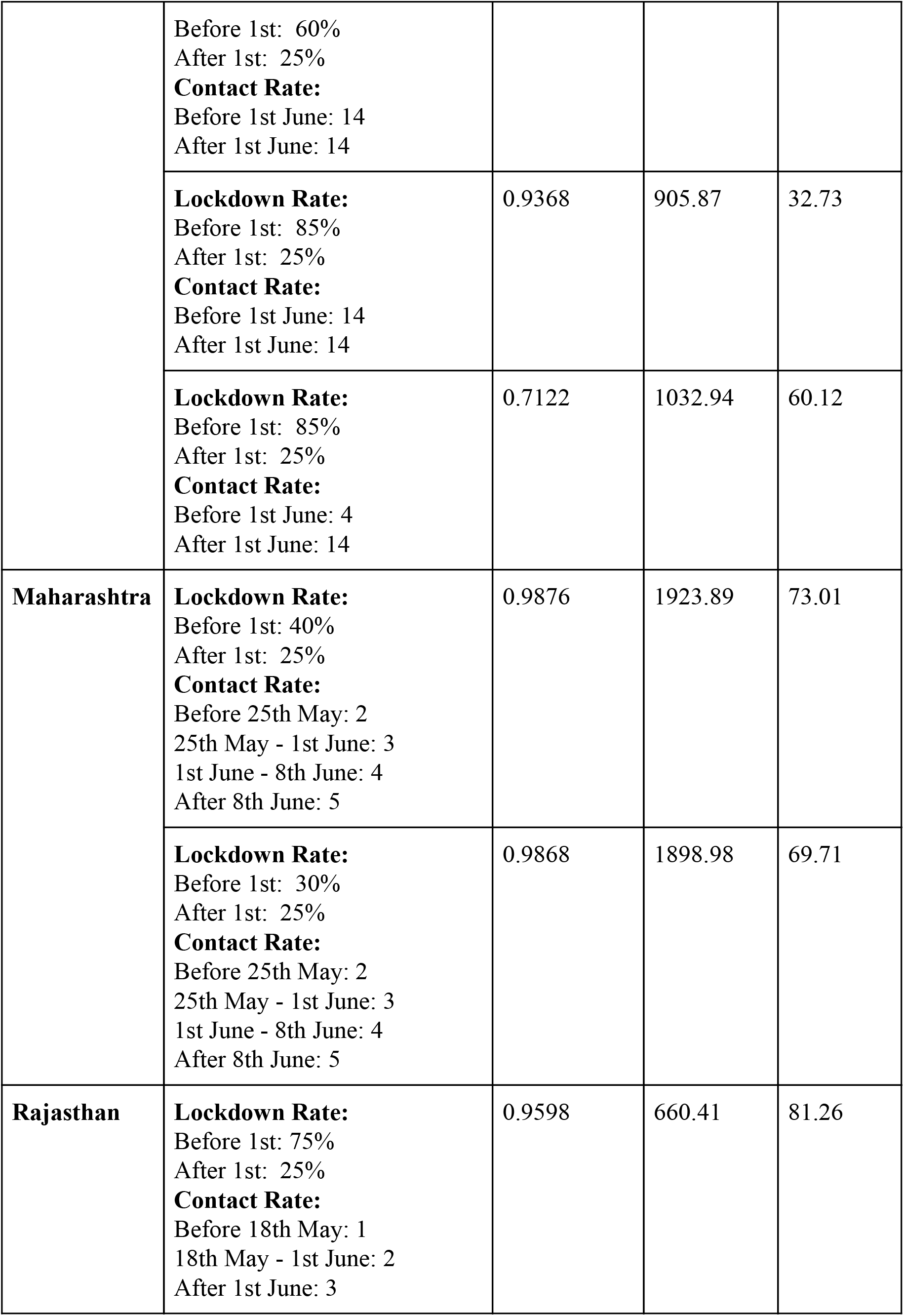
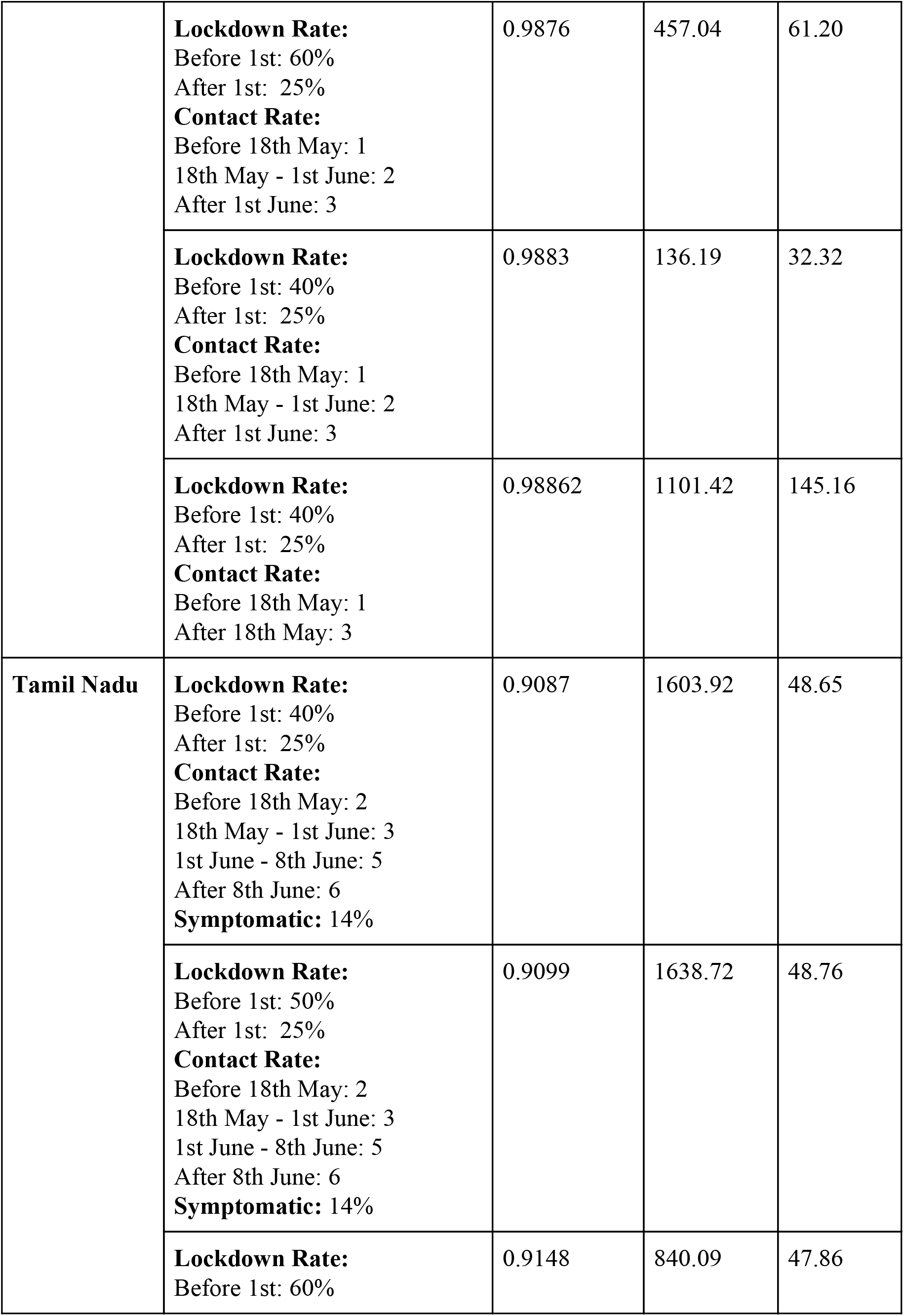
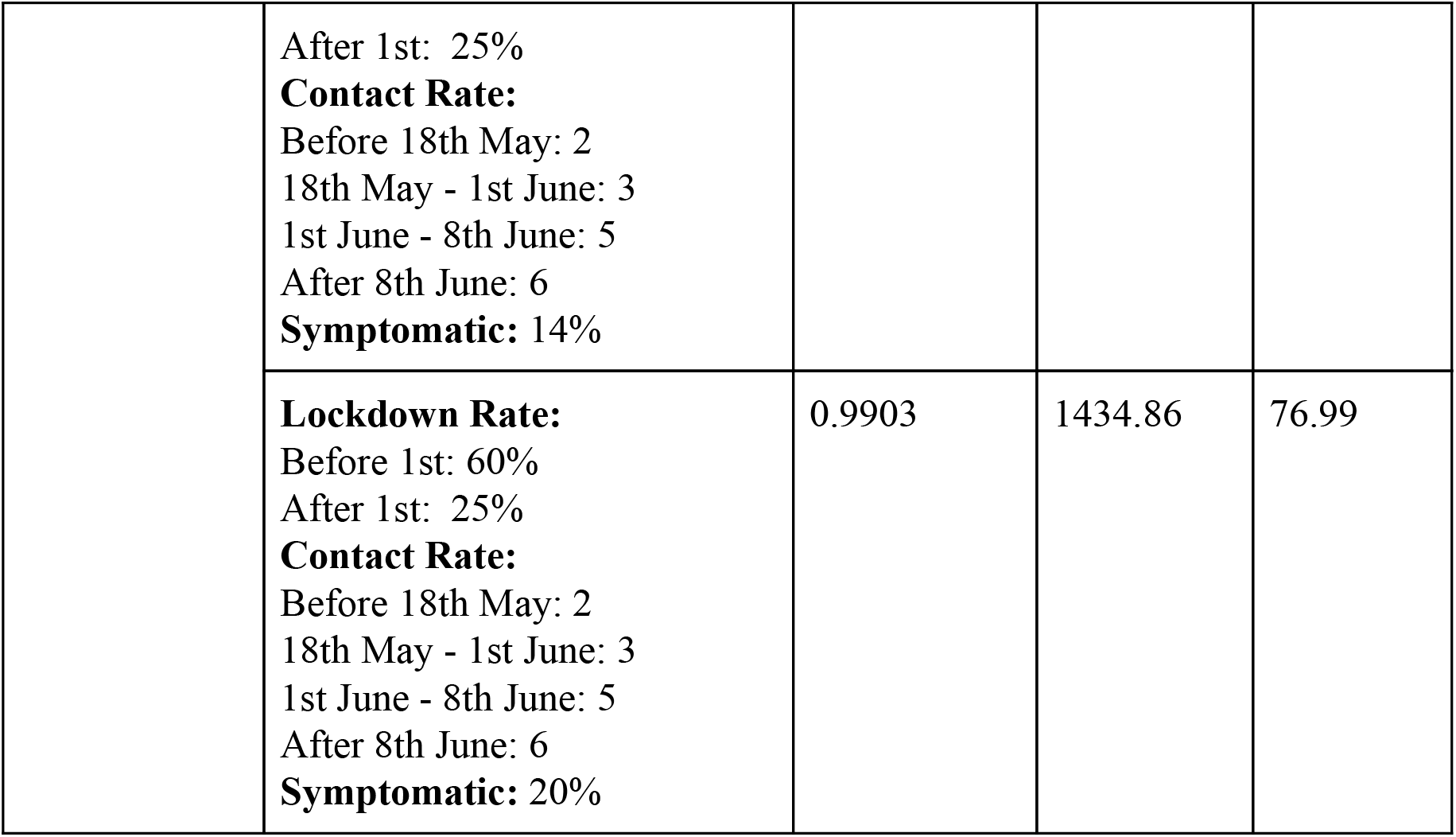
List of parameter combinations evaluated for next 30 days per-day predictions for each state.

| State | Scenario | R<sup>2</sup> | RMSE | MAPE |
| --- | --- | --- | --- | --- |
| **Gujarat** | **Lockdown Rate:**<br>Before 1st: 60%<br>After 1st: 25%<br>**Contact Rate:**<br>Before 18th May: 2<br>18th May - 1st June: 4<br>1st June - 8th June: 6<br>After 8th June: 8 | 0.9925 | 813.95 | 47.83 |
| **Madhya Pradesh** | **Lockdown Rate:**<br>Before 1st: 60%<br>After 1st: 25%<br>**Contact Rate:**<br>Before 18th May: 1<br>18th May - 1st June: 2<br>1st June - 8th June: 3<br>After 8th June: 4 | 0.9926 | 89.98 | 27.51 |
| **Delhi** | **Lockdown Rate:** | 0.9242 | 12390.63 | 418.91 |
|  | Before 1st: 60%<br>After 1st: 25%<br>**Contact Rate:**<br>Before 1st June: 14<br>After 1st June: 14 |  |  |  |
|  | **Lockdown Rate:**<br>Before 1st: 85%<br>After 1st: 25%<br>**Contact Rate:**<br>Before 1st June: 14<br>After 1st June: 14 | 0.9368 | 905.87 | 32.73 |
|  | **Lockdown Rate:**<br>Before 1st: 85%<br>After 1st: 25%<br>**Contact Rate:**<br>Before 1st June: 4<br>After 1st June: 14 | 0.7122 | 1032.94 | 60.12 |
| **Maharashtra** | **Lockdown Rate:**<br>Before 1st: 40%<br>After 1st: 25%<br>**Contact Rate:**<br>Before 25th May: 2<br>25th May - 1st June: 3<br>1st June - 8th June: 4<br>After 8th June: 5 | 0.9876 | 1923.89 | 73.01 |
|  | **Lockdown Rate:**<br>Before 1st: 30%<br>After 1st: 25%<br>**Contact Rate:**<br>Before 25th May: 2<br>25th May - 1st June: 3<br>1st June - 8th June: 4<br>After 8th June: 5 | 0.9868 | 1898.98 | 69.71 |
| **Rajasthan** | **Lockdown Rate:**<br>Before 1st: 75%<br>After 1st: 25%<br>**Contact Rate:**<br>Before 18th May: 1<br>18th May - 1st June: 2<br>After 1st June: 3 | 0.9598 | 660.41 | 81.26 |
|  | **Lockdown Rate:**<br>Before 1st: 60%<br>After 1st: 25%<br>**Contact Rate:**<br>Before 18th May: 1<br>18th May - 1st June: 2<br>After 1st June: 3 | 0.9876 | 457.04 | 61.20 |
|  | **Lockdown Rate:**<br>Before 1st: 40%<br>After 1st: 25%<br>**Contact Rate:**<br>Before 18th May: 1<br>18th May - 1st June: 2<br>After 1st June: 3 | 0.9883 | 136.19 | 32.32 |
|  | **Lockdown Rate:**<br>Before 1st: 40%<br>After 1st: 25%<br>**Contact Rate:**<br>Before 18th May: 1<br>After 18th May: 3 | 0.98862 | 1101.42 | 145.16 |
| Tamil Nadu | **Lockdown Rate:**<br>Before 1st: 40%<br>After 1st: 25%<br>**Contact Rate:**<br>Before 18th May: 2<br>18th May - 1st June: 3<br>1st June - 8th June: 5<br>After 8th June: 6<br>**Symptomatic:** 14% | 0.9087 | 1603.92 | 48.65 |
|  | **Lockdown Rate:**<br>Before 1st: 50%<br>After 1st: 25%<br>**Contact Rate:**<br>Before 18th May: 2<br>18th May - 1st June: 3<br>1st June - 8th June: 5<br>After 8th June: 6<br>**Symptomatic:** 14% | 0.9099 | 1638.72 | 48.76 |
|  | **Lockdown Rate:**<br>Before 1st: 60% | 0.9148 | 840.09 | 47.86 |
|  | After 1st: 25%<br>**Contact Rate:**<br>Before 18th May: 2<br>18th May - 1st June: 3<br>1st June - 8th June: 5<br>After 8th June: 6<br>**Symptomatic:** 14% |  |  |  |
|  | **Lockdown Rate:**<br>Before 1st: 60%<br>After 1st: 25%<br>**Contact Rate:**<br>Before 18th May: 2<br>18th May - 1st June: 3<br>1st June - 8th June: 5<br>After 8th June: 6<br>**Symptomatic:** 20% | 0.9903 | 1434.86 | 76.99 |

### 3. Contact Rate estimation equation

The non-linear scaling function focuses on the population density, disease spread, distance-based transmission to capture spatial distribution and the movement of population in an area. The equations used are given as follows:

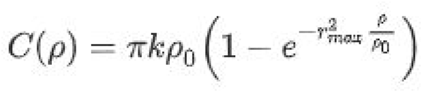

We use the following set of parameters in this scaling function:

**Supplementary Table 2:**
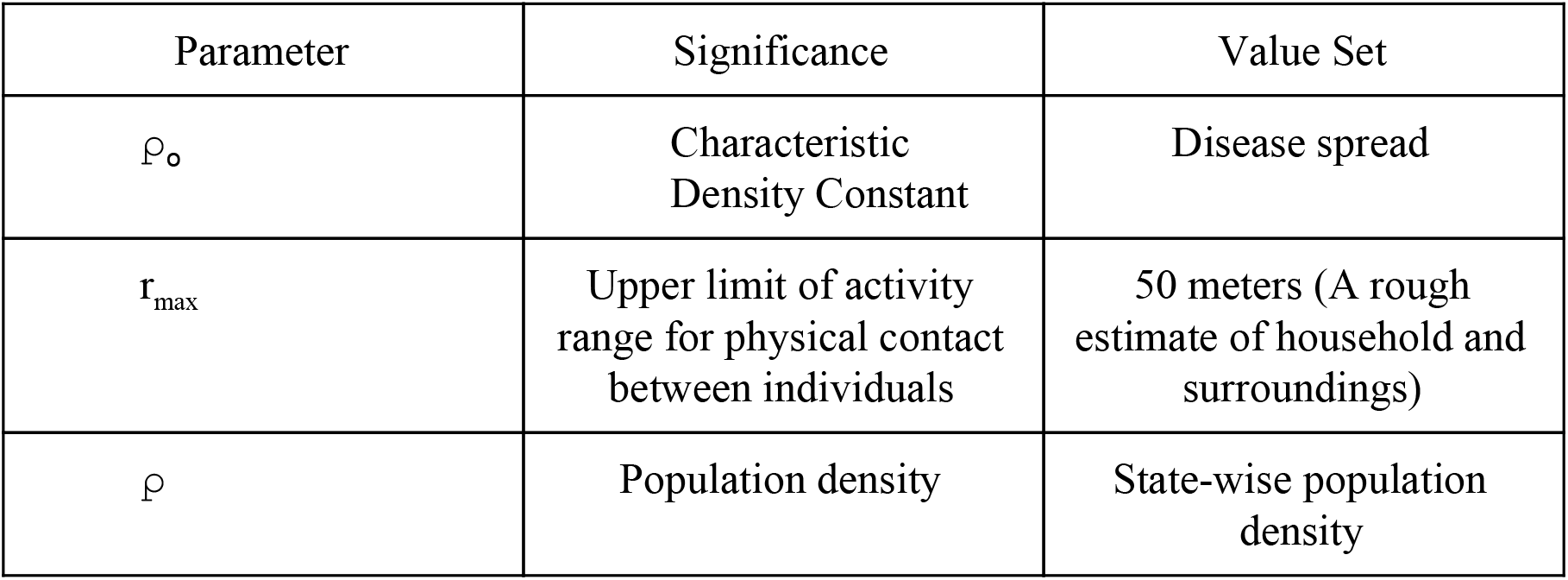
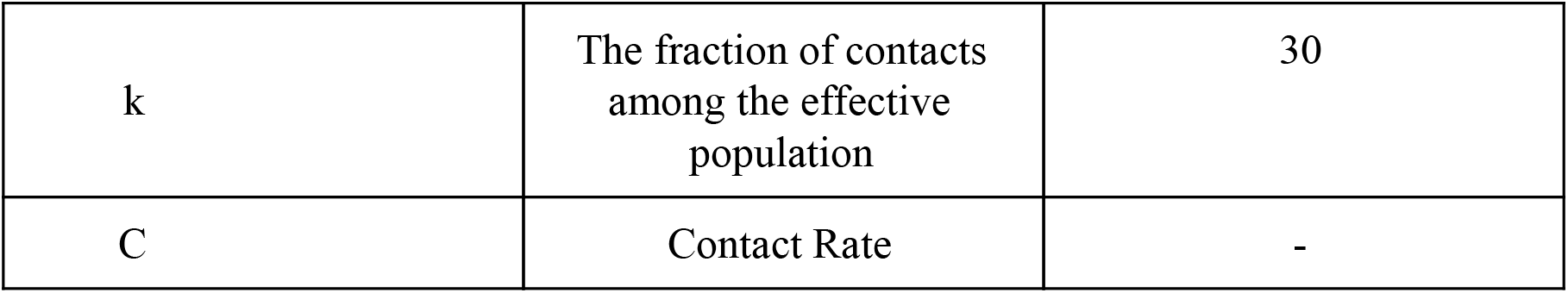
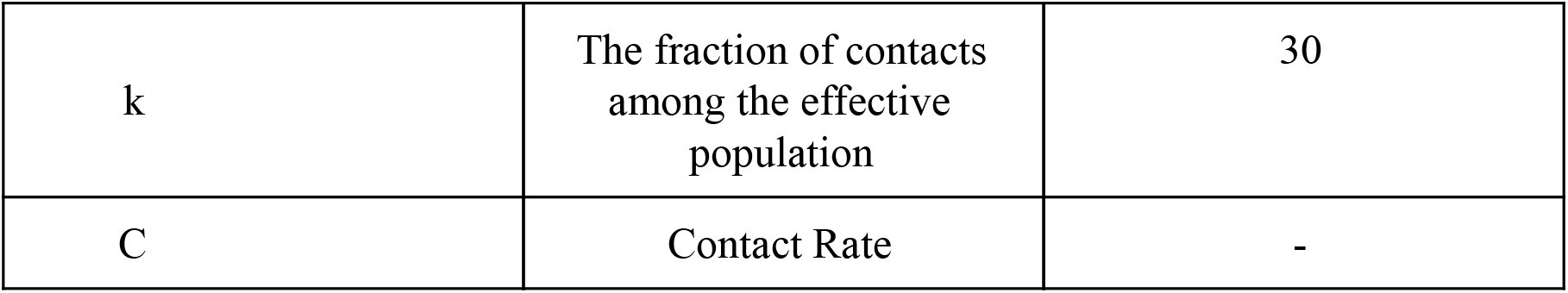
Different parameters used in the contact rate scaling equation.

| Parameter | Significance | Value Set |
| --- | --- | --- |
| $\rho_0$ | Characteristic Density Constant | Disease spread |
| $r_{\max}$ | Upper limit of activity range for physical contact between individuals | 50 meters (A rough estimate of household and surroundings) |
| $\rho$ | Population density | State-wise population density |
| k | The fraction of contacts among the effective population | 30 |
| C | Contact Rate | - |

**Supplementary Figure 3:**
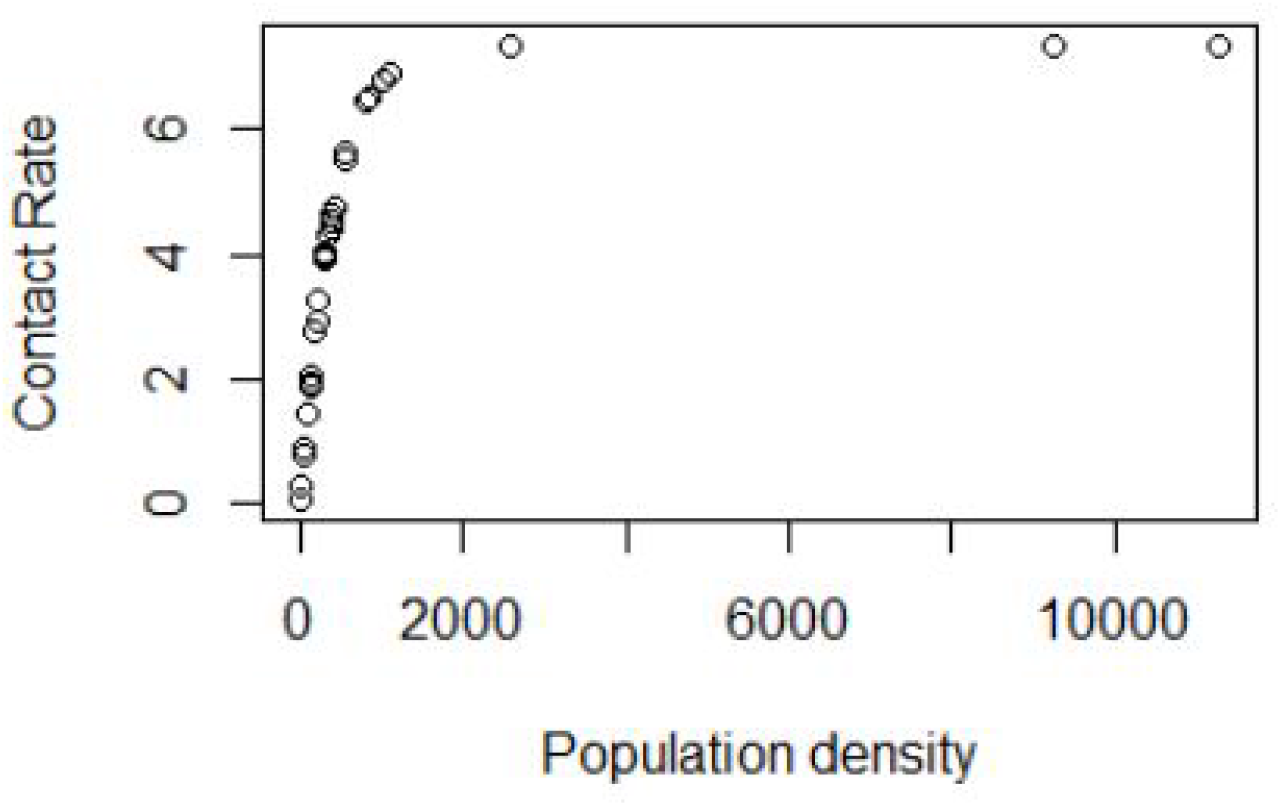
State-wise population density versus contact rate simulation.

## Compartmental Models

Compartmental models are differential equation models used to understand disease dynamics. Here we used the SIR and SEIR Model where (S for Susceptibility, E for Exposure, I for infectious, R for Removed/Recovery).

### SIR Model

In SIR model we divide the entire population N into three blocks S, I, R i.e. N= S+I+R. This model assumes that the force of infection in the present depends on the number of infectious agents till that point of time and hence estimates the rate of flow of the individuals between the compartments.

This is described by the following system of ordinary differential equations (ODEs):

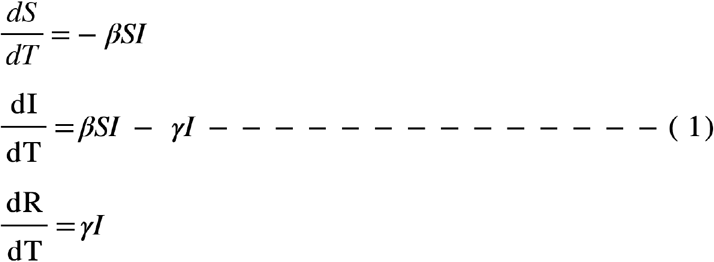

In equation (1) **β** controls how quickly susceptible population move to infectious and similarly ***γ*** controls how quickly infectious population move to recovery. and the ratio of **β** and ***γ*** is defined as Reproductive No.

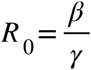

### SEIR Model

In the SEIR Model term E includes the latent phase of covid where individual is infected but not infectious. In the SEIR model we divide the total population N into 4 compartments S,E,I,R. in this model we include the parameter *σ*. threshold *σ* is identified which determines the outcome of the disease; if *σ* − 1, the infected fraction of the population disappears so the disease dies out, while if *σ* >1, the infected fraction persists and a unique endemic equilibrium state is shown, under a mild restriction on the parameters, to be globally asymptotically stable in the interior of the feasible region. i.e. N = S+E+I+R]. All four blocks can be modeled in by following equations

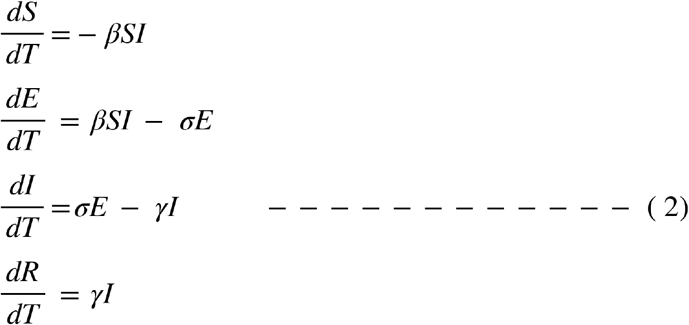

Latency in the disease (delayed start of the individual’s disease) is a parameter controlling this delay in the model. Inclusion of latency period in SEIR does not influence the Reproductive no (R0).

### Dashboard

An interactive dashboard http://covision.tavlab.iiitd.edu.in/ has been hosted as a web-server for the war level monitoring of the covid19 pandemic in India in public domain.

**Supplementary Figure 4:**
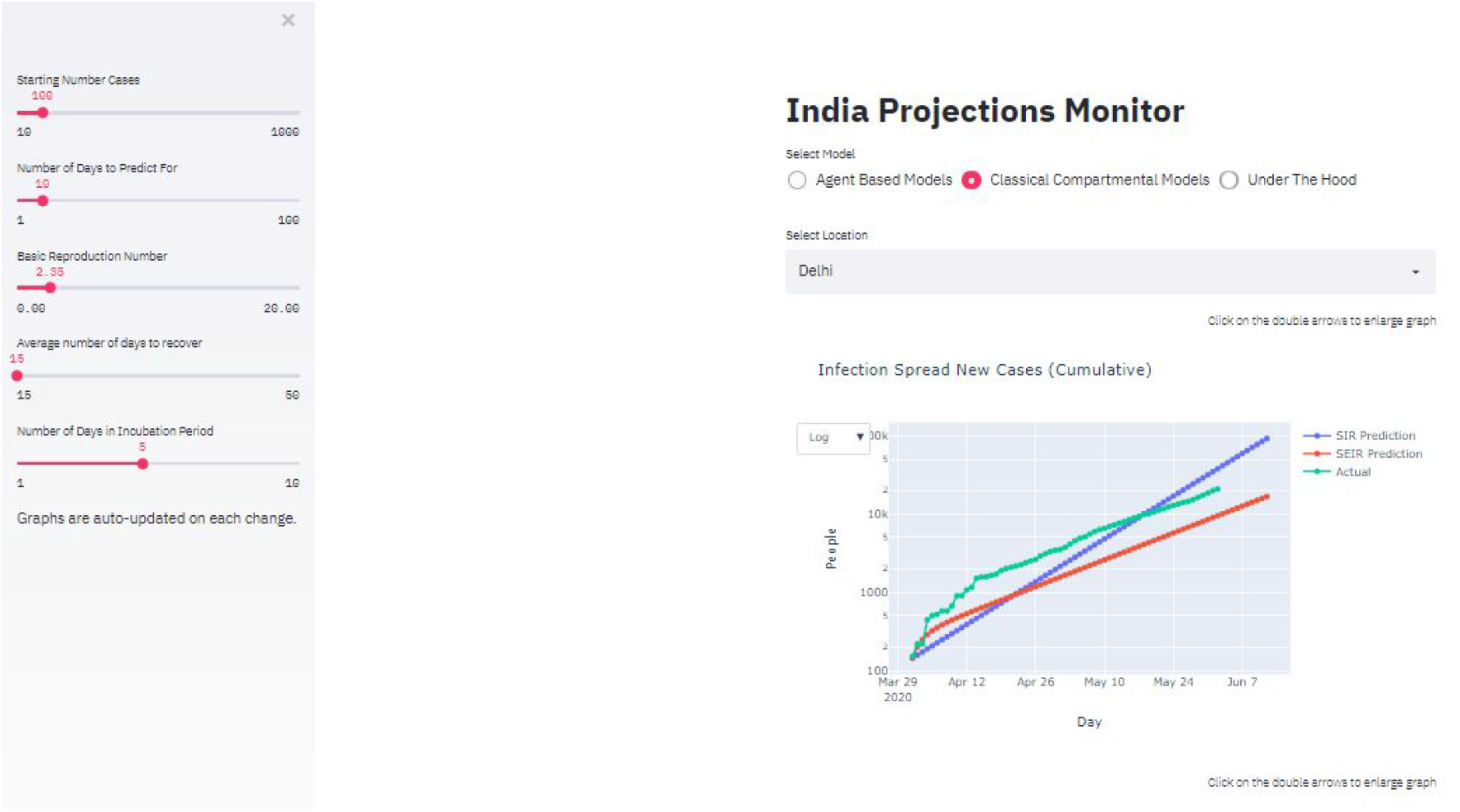
Interactive dashboard for Indian state-wise projections using different models and scenarios.

## References

1. Jay Naresh Dhanwant and V. Ramanathan. “Forecasting COVID 19 growth in India using the Susceptible-Infected-Recovered (S.I.R) model.”

2. Rajesh Ranjan. “Predictions For COVID-19 Outbreak In India Using Epidemiological Models.”

3. Singh and Adhikari. “Age-structured impact of social distancing on the COVID-19 epidemic in India.”

4. Venkatramanan et al “Using data-driven agent-based models for forecasting emerging infectious diseases.” ScienceDirect

5. Tuotino et al “An agent-based epidemic model REINA for COVID-19 to identify destructive policies” preprint MedaRxiv.

6. https://www.mohfw.gov.in/

7. https://data.gov.in/

8. https://web.archive.org/web/20200114212849/http://censusindia.gov.in/

9. Emrich, Stefan, Sergej Suslov, and Florian Judex. “Fully agent based modelling of epidemic spread using AnyLogic.” Proc. eurosim. 2007.

10. httPs://www.sciencedirect.com/science/article/pii/S0025556413001235?via%3Dihub

11. Soetaert, Karline ER, Thomas Petzoldt, and R. Woodrow Setzer. “Solving differential equations in R: package deSolve.” Journal of Statistical Software 33 (2010).

12. WHO report https://www.who.int/docs/default-source/coronaviruse/who-china-joint-mission-on-covid-19-final-report.pdf#:~:text=Using%20available%20preliminary%20data%2C.severe%20or%20critical%20disease.

13. JM, Rodriguez Llanes, M. G. Pedersen, and M. Meneghini. “Confronting COVID-19: Surging critical care capacity in Italy.” (2020).

14. Xie, Tong, Guan, Du, Haibu, Arthur S,. “Critical care crisis and some recommendations during Covid - 19 epidemic in China.” Intensive Care Medicine, SpringerLink (2020)

15. Phua, Weng, Ling, Egi, Ling, Divatia, “Intensive care management of coronavirus diseases 2019 (COVID 19): challenges and recommendations”

16. Vincent, Jean-Louis, and Fabio S. Taccone. “Understanding pathways to death in patients with COVID-19.” The Lancet Respiratory Medicine (2020).

17. https://www.who.int/docs/default-source/coronaviruse/situation-reports/20200306-sitrep-46-covid-19.pdf?sfvrsn=96b04adf_4

18. Unlock 1.0 Guidelines: https://mha.gov.in/sites/default/files/MHAD0LrDt_3052020.pdf

19. Population Data https://www.worldometers.info/world-population/population-by-country/

20. Holmdahl et al “Wrong but Useful — What Covid-19 Epidemiologic Models Can and Cannot Tell Us” The New England Journal of Medicine (2020)

21. Beretta, Edoardo, and Yasuhiro Takeuchi. “Global stability of an SIR epidemic model with time delays.” Journal of mathematical biology 33.3 (1995): 250-260.

22. Li, Michael Y., et al. “Global dynamics of a SEIR model with varying total population size.” Mathematical biosciences 160.2 (1999): 191-213.

23. Zhu, Caillou, et al. “Algorithm 778: L-BFGS-B: Fortran subroutines for large-scale bound-constrained optimization.” ACM Transactions on Mathematical Software (TOMS) 23.4 (1997): 550-560.

24. Kermack and McKendrick. “A contribution to the mathematical theory of epidemics.”

